# Associations of alcohol consumption with left atrial morphology and function in a population at high cardiovascular risk

**DOI:** 10.1101/2023.04.27.23289215

**Authors:** Aniqa B. Alam, Estefania Toledo-Atucha, Dora Romaguera, Angel M. Alonso-Gómez, Miguel A. Martínez-Gonzalez, Lucas Tojal-Sierra, Marta Noris Mora, Caterina Mas-Llado, Linzi Li, Ines Gonzalez-Casanova, Jordi Salas-Salvadó, Montserrat Fitó, Alvaro Alonso

## Abstract

**Background:** Excessive alcohol consumption has been associated with increased risk of atrial fibrillation, though the underlying mechanisms remain unclear. An enlarged left atrium and impaired left atrial function may lead to atrial fibrillation. The association of alcohol consumption with structural and functional left atrial measures, however, has received limited attention.

**Methods:** We studied a subset of 515 participants from the PREDIMED-Plus trial, a Spanish ongoing, multi-center randomized trial testing an intensive life weight loss intervention (ILI) with an energy reduced Mediterranean diet and physical activity promotion versus usual care and *ad libitum* Mediterranean diet recommendations for the primary prevention of cardiovascular disease in adults with the metabolic syndrome. Participants underwent transthoracic echocardiography (TTE) at baseline, year 3, and year 5 of the study. Outcomes of interest included volumetric (volume index) and functional (reservoir, conduit, and contractile strains) measures of the left atrium. Alcohol consumption was calculated through the use of validated food frequency questionnaires repeatedly collected every year and presented as drinks consumed per day (1 drink = 14 grams of alcohol). Confounder-adjusted multiple linear regression and mixed models were used to estimate the association of alcohol consumption (at baseline and over 5-years of follow-up) with left atrial volume and function at baseline and over the 5-year follow-up.

**Results:** Mean alcohol consumption among 515 eligible participants (mean age: 65 years, 40% female) was 1.6 drinks/day. Cross-sectionally, higher alcohol consumption (modeled in 1 drink/day increases) was associated with larger left atrial volume (0.6 ml/m^2^, 95% CI 0.2, 1.1) and lower left atrial reservoir and contractile strain (-0.4%, 95% CI -0.8, 0.0, and -0.4, 95% CI -0.7, -0.1). Baseline alcohol consumption was not associated with changes in left atrial structure and function, but an increase in alcohol consumption (per 1 drink/day increase) during follow-up was associated with left atrial enlargement (0.7 mL/m^2^, 95% CI 0.2, 1.2) and, to a lesser extent, left atrial strain worsening (-0.3%, 95% CI -0.8, 0.2) over 5 years.

**Conclusion:** In a population at high risk of cardiovascular disease, an increase of alcohol consumption was associated with left atrial volume enlargement and worsening atrial function, suggesting mechanistic information about potential pathways linking excessive alcohol consumption with atrial fibrillation risk.

## INTRODUCTION

Alcohol consumption is widespread in Western countries. Excessive alcohol consumption is associated with cardiovascular pathologies such as cardiomyopathy,^1^ stroke,^2,3^ myocardial infarction,^4,5^ and atrial fibrillation (AF).^6^ Even moderate drinking has been associated with incident AF,^7,8^ though to a lesser extent compared to heavy drinking, suggesting a dose-response relationship. It is important to note, however, that the classification of drinking severity varies across studies.

The mechanisms linking alcohol consumption and AF are not well-characterized. Previous studies have shown that increased left atrial diameter and dysfunction are associated with both excessive alcohol consumption and AF, with nearly 24% of the association of alcohol and AF being explained by increased left atrial diameter.^9,10^ Prior studies, however, did not consider more relevant measures of left atrial structure (such as left atrial volume), were performed in very small samples, or were merely cross-sectional. Furthermore, the effects of different types of alcohol on left atrial structure and function or AF risk are not well-studied.^7^ Thus, we propose examining the association of alcohol consumption, as well as consumption of alcohol from different sources, with volumetric and functional measures of the left atrium, both cross-sectionally and longitudinally.

## METHODS

### Study population

The PREDIMED-Plus study is an ongoing, randomized, and controlled lifestyle intervention trial being conducted in several centers throughout Spain with the primary aim of testing an intensive lifestyle intervention focusing on weight loss in the context of increasing adherence to an energy-reduced Mediterranean diet (MedDiet) and physical activity among overweight and obese individuals with the metabolic syndrome.^11^ Participants in the control group received a nutritional intervention to foster their adherence to the MedDiet with no total energy reduction nor advice on physical activity. This study’s protocol has been reviewed and approved by Institutional Review Boards at all participating institutions. All participants have provided written informed consent to be part of the study.

A subgroup of 566 participants recruited from three sites of the PREDIMED-Plus cohort (Navarra, Balearic Islands and Vitoria) who agreed to be included in the study underwent two-dimensional transthoracic echocardiography (TTE) at baseline, year 3, and year 5 of the study. After excluding those missing left atrium (LA) measurements at baseline (N = 51) and those with AF at baseline (N = 12), 503 participants were included in the present analysis (**Figure 1**).

**Figure 1.**
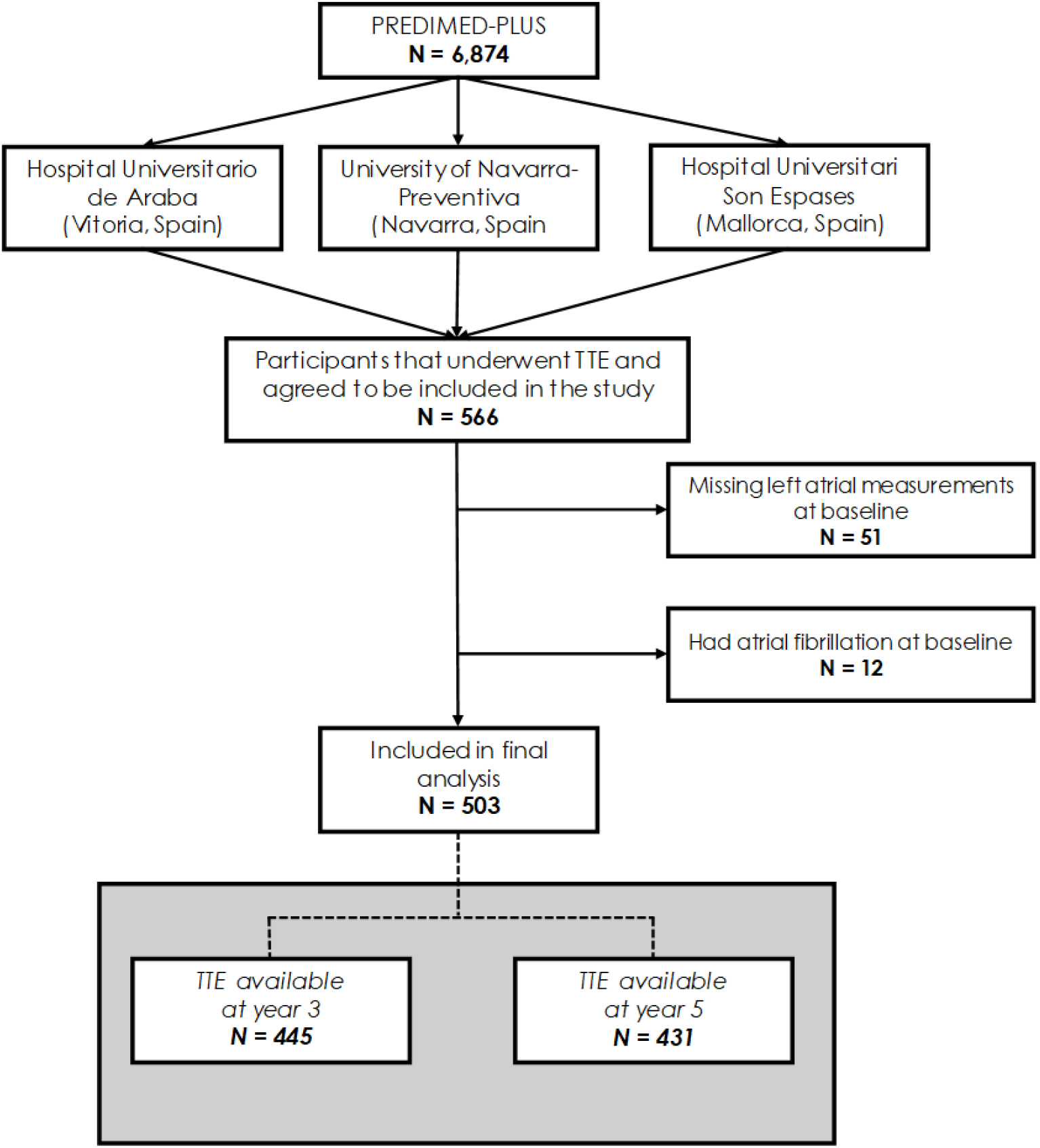
Flowchart of PREDIMED-Plus participants included into study.

### Left atrial structure and volume

Measurements at all sites were performed by trained personnel using GE Vivid machines, with studies sent to a central laboratory for reading by two dedicated cardiologists. Outcomes of interest included structural (left atrial volume index, LAVi) and functional (reservoir, conduit, and contractile atrial strains) measures of the left atrium. Maximal left atrial volume was measured before mitral valve opening using two-dimensional speckle tracking analysis of the left atrium, with four- and two-chamber apical views analyzed using AFI LA (GE EchoPAC),^12^ and indexed to body surface area to calculate LAVi. Left atrial strain indices were calculated with speckle-tracking software with an autostrain algorithm (GE Echopac). Detailed descriptions of the imaging process can be found elsewhere.^13^

### Alcohol consumption

Alcohol consumption was calculated with a validated 143-item semiquantitative food frequency questionnaire (FFQ) which participants filled out at baseline and every yearly follow-up visit.^14^ Specific categories of alcohol included: wine (red wine [aged and young], other wine [white, rosé, muscatel, cava]), beer, and spirits (liquor, whiskey). Since the pure alcohol content varies depending on the type of drink, standard sizes of each type of beverage in the FFQ were as follows: 1 glass of wine, 100 cc, except for muscatel in 50 cc; 1 bottle of beer, 330 cc; 1 shot of liquor or other spirits, 50 cc. In this analysis, one standard drink was equivalent to 14 grams of pure alcohol. For estimated alcohol intake, the intra-class correlation between the FFQ and four 3-day dietary records was 0.82.^14^

### Covariates

Education and marital status served as markers of socioeconomic status in this study. Physical activity was assessed at baseline in metabolic equivalents of task (METs) in minutes per day.^15^ Body-mass index (BMI) was calculated from weight and height at baseline. Smoking status was self-reported at baseline. Diet adherence was calculated using a 17-item questionnaire evaluating adherence to an energy-reduced MedDiet,^16^ but was recalculated to exclude alcohol intake to avoid redundancy. Systolic and diastolic blood pressure were taken 3 times at every visit and then averaged. Diabetes was based on self-reported information at baseline, use of antidiabetic medication, and measures of fasting blood glucose and glycosylated hemoglobin. The presence of depressive symptoms was evaluated using the 21-question Beck Depression Inventory.

Assignment to the study intervention group was also taken into consideration.

### Statistical analysis

Analyses were conducted using a dataset with follow-up data up to 5 years generated on August 10, 2022. Multivariable linear regression was used to estimate the cross-sectional association of baseline alcohol consumption with baseline LA measures. Mixed models were used to estimate longitudinal associations of baseline alcohol consumption with changes in LA measures from baseline to year 3 to year 5, with time being modeled continuously. We also evaluated the impact of changes in drinking amount over the course of the study by using the difference in alcohol consumption from baseline to year 5 as a predictor of changes in LA measures from baseline to year 5. All analyses were repeated to also produce estimates for each type of alcohol. Alcohol consumption was categorized into 0 (reference group), 1, 2-3, ≥ 4 drinks consumed per day.

Models were also ran using alcohol consumption as a continuous variable (modeled in drinks/day).

All models were initially adjusted for age, sex, education, and intervention group. They were further adjusted for marital status, smoking, physical activity, height, BMI, systolic and diastolic blood pressure, diabetes, depression, adherence to the energy-reduced MedDiet, and intervention group. Longitudinal analysis also considered interaction of all covariates with time. Changes in these covariates, however, may also influence LA structure and functioning, so differences from baseline to year 5 for the following covariates were also incorporated in our models: smoking, physical activity, BMI, systolic and diastolic blood pressure, diabetes, depression, diet adherence.

All analyses were performed using SAS 9.4 (Cary, NC; SAS Institute Inc.).

## RESULTS

Of 503 participants included in this study [mean age (SD): 65.1 (4.9) years], 40% were female and 51% were randomized to the intervention group. Heavier drinkers at baseline were younger, more educated, and more likely to be male than their non-drinking counterparts (**Table 1**).

**Table 1.** Cohort descriptives, by daily drinking frequency. 1 drink = 14 grams of alcohol.

| Characteristic | 0 drinks<br>(N = 99) | 1 drink/day<br>(N = 228) | 2-3 drinks/day<br>(N = 105) | ≥ 4 drinks/day<br>(N = 71) |
| --- | --- | --- | --- | --- |
| Intervention group, % | 54 | 51 | 48 | 52 |
| Age (years) | 66 (4) | 65 (5) | 65 (5) | 64 (5) |
| Female, % | 71 | 50 | 14 | 4 |
| Education, % |  |  |  |  |
| College | 11 | 13 | 16 | 20 |
| Technical school | 4 | 5 | 9 | 6 |
| High school | 25 | 28 | 30 | 49 |
| Primary school | 60 | 54 | 46 | 25 |
| Marital status, % |  |  |  |  |
| Single | 5 | 5 | 5 | 7 |
| Married | 73 | 75 | 82 | 86 |
| Widower | 15 | 12 | 8 | 3 |
| Divorced | 6 | 6 | 4 | 3 |
| Other | 1 | 1 | 2 | 1 |
| Height (cm) | 160 (9) | 163 (9) | 168 (8) | 170 (7) |
| Body-mass index (kg/m<sup>2</sup>) | 32.5 (3.6) | 32.2 (3.3) | 32.0 (3.0) | 32.3 (3.0) |
| Systolic blood pressure (mmHg) | 142 (17) | 140 (17) | 141 (16) | 143 (16) |
| Diastolic blood pressure (mmHg) | 77 (9) | 78 (10) | 81 (9) | 83 (10) |
| Smoking, % |  |  |  |  |
| Currently smoking | 5 | 10 | 9 | 13 |
| Former smoker (0-5 years ago) | 4 | 5 | 8 | 10 |
| Former smoker (>5 years ago) | 31 | 42 | 56 | 58 |
| Never smoker | 60 | 43 | 28 | 20 |
| Physical activity (MET minutes/week) | 2244 (1906) | 2398 (2066) | 3104 (2966) | 2471 (2163) |
| Diabetes, % | 43 | 29 | 18 | 20 |
| BECK depression inventory (0-63) | 9.3 (7.3) | 8.3 (7.3) | 7.9 (7.7) | 6.7 (5.6) |
| Diet adherence* (1-16) | 8.1 (2.7) | 7.6 (3.0) | 6.7 (2.9) | 5.5 (2.6) |
| Daily wine consumption (g/d) | 0 | 35 (34) | 170 (107) | 452 (181) |
| Daily beer consumption (g/d) | 0 | 49 (74) | 192 (245) | 280 (474) |
| Daily spirits consumption (g/day) | 0 | 1 (3) | 6 (10) | 13 (25) |
| *Left atrial outcomes* |  |  |  |  |
| LAVi, mL/m<sup>2</sup> | 22.6 (6.8) | 22.6 (7.2) | 22.0 (6.8) | 25.1 (6.9) |
| LA reservoir strain | 27.4 (6.5) | 27.6 (6.7) | 28.2 (6.8) | 27.3 (5.2) |
| LA conduit strain | 11.4 (3.7) | 12.0 (4.8) | 12.4 (4.7) | 11.8 (3.4) |
| LA contractile strain | 15.9 (5.0) | 15.6 (4.7) | 15.7 (4.4) | 15.5 (4.3) |

Presented as either mean (standard deviation) for continuous measures or as frequencies for categorical variables.
LA, left atrial. LAVi: left atrial volume index.
\*Sum of 16 questions concerning adherence to an energy reduced Mediterranean diet. Does not include alcohol intake.
1 drink: 1-14 grams. 2-3 drinks: 15-42 grams. 4 or more drinks: 43 grams and more

Echocardiographic measurements were available in 445 participants at year 3 (88%) and 431 participants at year 5 (86%) (**Figure 1**).

Cross-sectionally, modeling of alcohol consumption as a continuous variable showed larger LAVi with higher alcohol consumption (β: 0.6 mL/m^2^, 95% 0.2, 1.1 per 1 drink/day), after adjustment for demographic, lifestyle and clinical variables (**Table 2**). Considering categories of alcohol intake suggested a J-shaped association, with higher LAVi observed only at highest levels of alcohol consumption (**Table 2**). Higher baseline alcohol consumption showed linear associations with reduced left atrial reservoir strain and left atrial contractile strain, but not with left atrial conduit strain (**Table 2**). One drink/day was associated with reductions of -0.4% (95%CI -0.9, 0.0) of left atrial reservoir strain and -0.4% (95%CI -0.8, -0.1) of left atrial contractile strain.

**Table 2.** Multiple linear regression estimates of overall alcohol consumption with LA measures at baseline.

|  |  | **0 drinks** | **1 drink/day** | **2-3 drinks/day** | **≥ 4 drinks/day** | **Per 1-drink\***<br>**increase** |
| --- | --- | --- | --- | --- | --- | --- |
| LAVi, mL/m<sup>2</sup> | Model 1 | 0 (ref) | 0.1 (-1.6, 1.8) | -0.4 (-2.5, 1.7) | 2.7 (0.4, 5.1) | 0.7 (0.2, 1.1) |
|  | Model 2 | 0 (ref) | -0.1 (-1.8, 1.6) | -0.8 (-2.9, 1.4) | 2.3 (-0.1, 4.8) | 0.6 (0.2, 1.1) |
| Left atrial reservoir strain, % | Model 1 | 0 (ref) | -0.3 (-1.9, 1.2) | -0.5 (-2.4, 1.4) | -1.5 (-3.6, 0.6) | -0.4 (-0.8, 0.0) |
|  | Model 2 | 0 (ref) | -0.7 (-2.3, 0.8) | -1.0 (-3.0, 1.0) | -1.9 (-4.2, 0.3) | -0.4 (-0.9, 0.0) |
| Left atrial conduit strain, % | Model 1 | 0 (ref) | 0.4 (-0.6, 1.4) | 0.5 (-0.8, 1.8) | -0.1 (-1.5, 1.4) | -0.1 (-0.3, 0.2) |
|  | Model 2 | 0 (ref) | 0.5 (-0.5, 1.6) | 0.9 (-0.4, 2.2) | 0.5 (-1.0, 2.0) | 0.0 (-0.3, 0.3) |
| Left atrial contractile strain, % | Model 1 | 0 (ref) | -0.6 (-1.7, 0.5) | -1.1 (-2.4, 0.3) | -1.4 (-2.9, 0.2) | -0.3 (-0.6, 0.0) |
|  | Model 2 | 0 (ref) | -1.0 (-2.1, 0.2) | -1.6 (-3.1, -0.2) | -2.1 (-3.7, -0.5) | -0.4 (-0.8, -0.1) |

\*1 drink = 14 grams of alcohol
LAVi, left atrial volume index.
Model 1: adjusted for age, sex, education.
Model 2: adjusted for age, sex, education, marital status, smoking, physical activity, height, body mass index (BMI), systolic and diastolic blood pressure, diabetes, depression, and adherence to the energy-reduced Mediterranean diet.

Baseline alcohol consumption was not associated with changes in left atrial volume nor with strain measures (**Table 3**). However, changes in alcohol consumption over 5 years were associated with changes in left atrial structure and function. Increasing alcohol consumption by at least 1 drink per day was associated with 0.7 mL/m^2^ (95%CI: 0.2, 1.3) increases in LAVi compared to participants that did not change amount of alcohol consumption (**Table 4**).

**Table 3.** Mixed models estimates of overall alcohol consumption at baseline with change in LA measures from baseline to year 5.

|  |  | **0 drinks** | **1 drink/day** | **2-3 drinks/day** | **≥ 4 drinks/day** | **Per 1-drink\* increase** |
| --- | --- | --- | --- | --- | --- | --- |
| Change in LAVi, mL/m<sup>2</sup> | Model 1 | 0 (ref.) | -0.2 (-0.6, 0.1) | 0.1 (-0.3, 0.6) | 0.1 (-0.4, 0.6) | 0.1 (0.0, 0.2) |
|  | Model 2 | 0 (ref.) | -0.2 (-0.6, 0.2) | 0.1 (-0.3, 0.6) | 0.0 (-0.5, 0.5) | 0.0 (-0.1, 0.1) |
|  | Model 3 | 0 (ref.) | -0.2 (-0.6, 0.2) | 0.2 (-0.3, 0.7) | -0.1 (-0.7, 0.5) | 0.0 (-0.1, 0.1) |
| Change in LA reservoir strain, % | Model 1 | 0 (ref.) | 0.0 (-0.4, 0.3) | -0.1 (-0.6, 0.3) | 0.3 (-0.2, 0.8) | 0.1 (0.0, 0.2) |
|  | Model 2 | 0 (ref.) | 0.0 (-0.4, 0.3) | -0.2 (-0.7, 0.3) | 0.3 (-0.2, 0.8) | 0.1 (0.0, 0.2) |
|  | Model 3 | 0 (ref.) | 0.0 (-0.4, 0.4) | -0.1 (-0.6, 0.4) | 0.4 (-0.2, 0.9) | 0.1 (0.0, 0.2) |
| Change in LA conduit strain, % | Model 1 | 0 (ref.) | 0.0 (-0.3, 0.2) | -0.1 (-0.4, 0.2) | 0.2 (-0.2, 0.5) | 0.0 (0.0, 0.1) |
|  | Model 2 | 0 (ref.) | -0.1 (-0.3, 0.2) | -0.2 (-0.5, 0.1) | 0.1 (-0.3, 0.4) | 0.0 (0.0, 0.1) |
|  | Model 3 | 0 (ref.) | -0.2 (-0.4, 0.1) | -0.2 (-0.6, 0.1) | 0.1 (-0.3, 0.5) | 0.0 (0.0, 0.1) |
| Change in LA contractile strain, % | Model 1 | 0 (ref.) | 0.0 (-0.2, 0.3) | 0.0 (-0.3, 0.3) | 0.1 (-0.2, 0.5) | 0.0 (0.0, 0.1) |
|  | Model 2 | 0 (ref.) | 0.0 (-0.2, 0.3) | 0.0 (-0.3, 0.4) | 0.2 (-0.2, 0.6) | 0.1 (0.0, 0.1) |
|  | Model 3 | 0 (ref.) | 0.1 (-0.1, 0.4) | 0.1 (-0.2, 0.5) | 0.3 (-0.1, 0.7) | 0.0 (0.0, 0.1) |

\*1 drink = 14 grams of alcohol
LA, left atrial. LAVi, left atrial volume index.
Model 1: adjusted for age, sex, education, intervention group, and interaction of all covariates with time
Model 2: adjusted for age, sex, education, intervention group, marital status, smoking, physical activity, height, body mass index (BMI), systolic and diastolic blood pressure, diabetes, depression, adherence to the Mediterranean diet, and interaction of all covariates with time.
Model 3: adjusted for age, sex, education, intervention group, marital status, smoking, physical activity, height, body mass index (BMI), systolic and diastolic blood pressure, diabetes, depression, adherence to the Mediterranean diet, interaction of all covariates with time, and differences from baseline to year 5 for the following: smoking, physical activity, BMI, systolic and diastolic blood pressure, diabetes, depression, and adherence to the Mediterranean diet.

**Table 4.** Mixed models estimates of changes in alcohol consumption with change in LA measures from baseline to year 5.

|  |  | **Decreased consumption**<br>**(≥ 1 drinks/day)** | **No change**<br>**(-1 > drinks/day > 1)** | **Increased consumption**<br>**(≥ 1 drinks/day)** | **Per 1-drink\* increase** |
| --- | --- | --- | --- | --- | --- |
| Change in LAVi,<br>mL/m<sup>2</sup> | Model 1 | 0.1 (-0.2, 0.5) | 0 (Ref.) | 0.8 (0.3, 1.3) | 0.2 (0.0, 0.4) |
|  | Model 2 | 0.1 (-0.3, 0.4) | 0 (Ref.) | 0.7 (0.2, 1.3) | 0.2 (0.0, 0.3) |
|  | Model 3 | 0.1 (-0.3, 0.5) | 0 (Ref.) | 0.7 (0.2, 1.3) | 0.2 (0.0, 0.3) |
| Change in LA<br>reservoir strain, % | Model 1 | 0.0 (-0.4, 0.3) | 0 (Ref.) | -0.2 (-0.7, 0.3) | -0.1 (-0.2, 0.1) |
|  | Model 2 | -0.1 (-0.4, 0.3) | 0 (Ref.) | -0.3 (-0.8, 0.3) | -0.1 (-0.3, 0.1) |
|  | Model 3 | 0.1 (-0.3, 0.5) | 0 (Ref.) | -0.2 (-0.7, 0.3) | -0.1 (-0.3, 0.1) |
| Change in LA<br>conduit strain, % | Model 1 | 0.0 (-0.2, 0.3) | 0 (Ref.) | -0.1 (-0.4, 0.3) | 0.0 (-0.1, 0.1) |
|  | Model 2 | 0.0 (-0.2, 0.3) | 0 (Ref.) | -0.1 (-0.4, 0.3) | -0.1 (-0.2, 0.1) |
|  | Model 3 | 0.1 (-0.1, 0.4) | 0 (Ref.) | -0.1 (-0.4, 0.3) | -0.1 (-0.2, 0.1) |
| Change in LA<br>contractile strain,<br>% | Model 1 | -0.1 (-0.3, 0.2) | 0 (Ref.) | -0.1 (-0.5, 0.2) | -0.1 (-0.2, 0.1) |
|  | Model 2 | -0.1 (-0.3, 0.1) | 0 (Ref.) | -0.2 (-0.5, 0.2) | -0.1 (-0.2, 0.1) |
|  | Model 3 | 0.0 (-0.3, 0.3) | 0 (Ref.) | -0.1 (-0.5, 0.3) | -0.1 (-0.2, 0.1) |

\*1 drink = 14 grams of alcohol
LA, left atrial. LAVi, left atrial volume index.
Model 1: adjusted for age, sex, education, intervention group, and interaction of all covariates with time
Model 2: adjusted for age, sex, education, intervention group, marital status, smoking, physical activity, height, body mass index (BMI), systolic and diastolic blood pressure, diabetes, depression, adherence to the Mediterranean diet, and interaction of all covariates with time.
Model 3: adjusted for age, sex, education, intervention group, marital status, smoking, physical activity, height, body mass index (BMI), systolic and diastolic blood pressure, diabetes, depression, adherence to the Mediterranean diet, interaction of all covariates with time, and differences from baseline to year 5 for the following: smoking, physical activity, BMI, systolic and diastolic blood pressure, diabetes, depression, and adherence to the Mediterranean diet.

Increasing alcohol consumption from baseline to year 5 was also non-significantly associated with worsening left atrial function, as determined by left atrial strain measurements (**Table 4**). Analyses modeling change in alcohol intake as a spline variable once again suggested a J-shaped association, with increases in LAVi observed only with exposure to heavy alcohol consumption (**Table 4** and **Figure 2)**. The associations of change in alcohol intake (when modeled as a spline variable) with strain measures were mostly linear (**Figure 2**).

**Figure 2.**
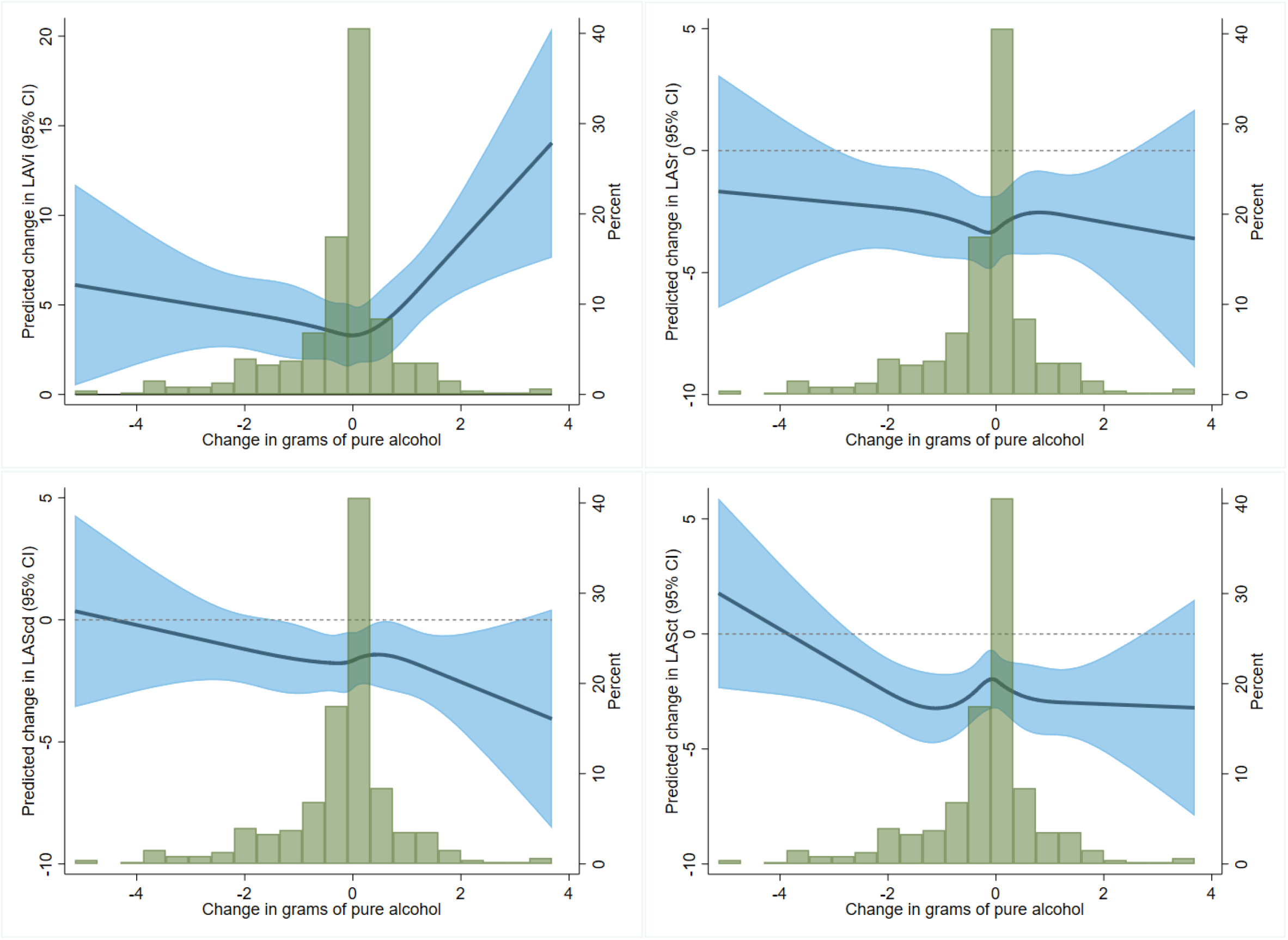
Associations of change in alcohol over 5 years modeled as restricted cubic splines with change in left atrial measures, using the median change in alcohol consumption as the reference point. Top left to right: left atrial volume, left atrial reservoir strain. Bottom left to right: left atrial conduit strain, left atrial contractile strain. Models adjusted for age, sex, education, and intervention group.

Heavy wine consumption was associated with significantly elevated LAVi at baseline (β: 2.6 mL/m^2^, 95% CI: 0.3, 4.8, comparing ≥ 4 drinks/day to 0 drinks), but no such associations were present with beer or spirits (**Supplemental Table 1)**. Heavy wine consumption was also associated with poorer reservoir (β: -2.1%, 95% CI: -4.1, 0.0) and contractile strain (β: -1.5%, 95% CI: -3.0, 0.0) at baseline. The different types of alcoholic beverage did not show any association with changes in left atrial measures from baseline to year 5 (**Supplemental Table 2**).

Higher alcohol consumption was associated with larger left atrial volumes (β: 3.7 mL/m^2^, 95% CI: 0.6, 6.8) and poorer contractile strains (β: -2.3%, 95% CI: -4.4, -0.2) at baseline in men, but not in women (**Supplemental Table 3**). There was, however, no evidence of sex-interaction [p-value = 0.76 (volume) and 0.20 (contractile)]. Once again, there was no evidence of longitudinal associations of alcohol consumption and changes in left atrial measures when analyses were stratified by sex (**Supplemental Table 4**).

Participants were also stratified by drinking intensity at baseline. Participants who began with 0-1 drinks per day and progressed to 2 or more drinks per day by year 5 had higher volumes (β: 0.7 mL/m^2^, 95% CI: 0.1, 1.3) and worse reservoir strains (β: -0.6%, 95% CI: -1.2, 0.0) compared to those who remained light drinkers (**Supplemental Table 5**). Participants who began with 2 or more drinks per day and reduced their intake to 0-1 drinks per day by year 5 had lower volumes, but also worse strain function compared to those who remained heavy drinkers; these estimates, however, were not significant.

## DISCUSSION

In a study of 503 Spanish adults at high cardiovascular risk we found that higher alcohol intake was cross-sectionally associated with LAVi following a J-shaped association and with impaired left atrial function. Baseline alcohol intake was not associated with changes in left atrial structure or function longitudinally over a 5-year period. However, increases in alcohol consumption during the study period were associated with increases in LAVi and, to a lesser extent, reductions in left atrial strain.

Similar findings were present in the community-based Atherosclerosis Risk in Communities Study, which reported higher left atrial size associated with higher alcohol intake in a cross-sectional analysis,^17^ and in the Framingham Heart Study, where higher alcohol intake was associated with higher left atrial size, but not with change in diameter over time.^9^ The authors suggested alcohol may increase left atrial size up to a certain point, after which the effect plateaus, which may explain the lack of longitudinal associations in this analysis. Furthermore, historical drinking behavior may have some influence on the progression of atrial remodeling and dilation, and as PREDIMED-Plus did not collect data on drinking patterns before the start of the trial, there may be some bias that we cannot account for in this analysis.

Studies conducted in populations with cardiovascular disease have also found associations between higher alcohol consumption and increased left atrial size or impaired left atrial function. In the Heart and Soul Study, among 601 patients with coronary heart disease, heavy alcohol consumption – based on the Alcohol Use Disorders Identification Test Consumption (AUDIT-C) test to identify alcohol use disorders – was associated with increases in left atrial volume over a 5-year period.^18^ Another study conducted in 160 patients with AF found higher left atrial volume and impaired left atrial function in those with higher alcohol consumption.^19^

Patterns of consumption are not typically static; it is important to account for changes in alcohol intake over time. Markers of left atrial health worsened in PREDIMED-Plus participants who increased their alcohol consumption after initial baseline assessments. While we did not find a reduction in intake to be particularly beneficial, other studies suggest that abstinence may reduce AF burden among frequent drinkers. In an open-label, randomized, controlled trial, Voskoboinik and colleagues reported significant reductions in alcohol intake in symptomatic AF patients resulted in reductions in AF events and time spent in AF.^20^ The availability of RCTs supporting these associations bolster the results of this present study by bridging the gap between morphological and functional changes in the myocardium and clinical presentation of atrial dysfunction. Unfortunately, there is limited additional evidence evaluating the impact of changes in alcohol consumption on markers of left atrial structure and function. This information is needed in order to better understand the role of alcohol on the risk of AF and to expand the findings of Voskoboinik et al to primary prevention settings with an anticipatory perspective.

Alcohol is the most commonly reported trigger for paroxysmal AF^21^ and is even thought to be the precipitator of nearly 35% of all new-onset cases of AF.^22^ Moreover, phenomena such as “holiday heart syndrome”, an alcohol-induced episode of AF, are common in emergency rooms.^23^ Alcohol may exercise its effects on left atrial substrate through both direct and indirect methods. Heavy alcohol consumption is an established cause of ventricular enlargement and cardiopathy,^24^ which itself may be on the pathway to atrial dysfunction.^25^ As the walls of the atria are thinner than the ventricles, the atria may be more susceptible to enlargement and myopathy due to alcohol consumption than the ventricles, resulting in elevated AF risk. Habitual consumption is also associated with common AF risk factors such as hypertension and oxidative stress.^25^

Stratifying our cohort by alcohol type and sex did not appear to impact our original findings. Wine-drinking was associated with higher left atrial volume and lower reservoir and contractile strains at baseline, but not with changes in left atrial measures by year 5 of the trial.

Consumption of beer and spirits did not seem to impact left atrial measures. Of note, in this older cohort from a Mediterranean country, wine was the main source of alcohol in the diet, with limited variability in the intake of beer or spirits. This lack of variability may explain the observed lack of associations. Wine consumption is part of the MedDiet, but its impact on AF and its related pathways is unclear. On the one hand, wine, particularly red wine, may have some antiarrhythmic benefits by way of the antioxidant resveratrol.^26,27^ For every cardioprotective finding, however, there is also evidence of proarrhythmic changes in response to moderate-to-high consumption of alcohol.^25,28^ It is important to note, however, that most of these studies to date rarely separate the effects of wine from other alcohol types in the context of AF risk. No clear sex differences were present in this study in the relationship between alcohol and left atrial measures. Though higher intake was associated with higher volume and lower contractile strains in men and not in women, there was no evidence of statistically significant interactions between sex and alcohol intake.

The main strengths of this analysis include the use of echocardiograms at multiple points within the trial, the use of a central site for reading of echocardiographic studies, the repeated dietary assessments, the availability of information on multiple potential confounders, and the excellent retention within the study, reducing the risk for informative censoring and subsequent selection bias. It is also important to take note of this analysis’s limitations. First, we do not have information on drinking behavior before the trial. Those who are life-long abstainers may be inherently different from those who abstained later on in life. Second, as alcohol intake in this study is based entirely on self-report, recall errors may be biasing our estimates. Finally, despite common protocols for the echocardiographic studies, within-person and between-person variability in echocardiographic image acquisition may obscure potential associations.

## CONCLUSION

In this well-characterized cohort of persons at high cardiovascular risk, we found evidence of morphological changes and impaired function of the left atrium associated with high or increasing alcohol intake. These results may provide some insight on the underlying mechanisms connecting alcohol and AF, and can contribute in informing recommendations related to alcohol consumption in persons at high risk of developing this arrhythmia.

## Data Availability

Interested researchers can request PREDIMED-Plus data directly from the study. Details for data acquisition are provided here: https://www.predimedplus.com/en/project/

