## Supplemental Tables for "Associations of alcohol consumption with left atrial morphology and function in a population at high cardiovascular risk"

Supplemental Table 1. Multiple linear regression estimates of each type of alcoholic beverage with LA measures at baseline.

| **Wine** | **0 drinks** | **1 drink/day** | **2-3 drinks/day** | **≥ 4 drinks/day** | **Per 1-drink increase** |
| --- | --- | --- | --- | --- | --- |
| Total LAVi | 0 (ref) | 0.6 (-1.1, 2.1) | 1.1 (-1.3, 3.4) | 2.6 (0.3, 4.8) | 0.7 (0.3, 1.1) |
| Left atrial reservoir strain | 0 (ref) | -1.1 (-2.6, 0.4) | -1.3 (-3.5, 0.9) | -2.1 (-4.1, 0.0) | -0.3 (-0.7, 0.1) |
| Left atrial conduit strain | 0 (ref) | -0.3 (-1.3, 0.7) | -0.1 (-1.6, 1.3) | -0.4 (-1.8, 0.9) | 0.0 (-0.3, 0.2) |
| Left atrial contractile strain | 0 (ref) | -0.6 (-1.7, 0.4) | -1.2 (-2.8, 0.3) | -1.5 (-3.0, 0.0) | -0.3 (-0.5, 0.0) |
| **Beer** | **0 drinks** | **1 drink/day** | **≥ 2 drinks/day** |  | **Per 1-drink increase** |
| Total LAVi | 0 (ref) | -0.1 (-1.5, 1.3) | 0.8 (-2.3, 3.8) |  | 0.2 (-0.7, 1.1) |
| Left atrial reservoir strain | 0 (ref) | -0.1 (-1.4, 1.2) | 0.3 (-2.5, 3.1) |  | -0.3 (-1.1, 0.5) |
| Left atrial conduit strain | 0 (ref) | 0.6 (-0.2, 1.5) | 2.0 (0.1, 3.9) |  | 0.4 (-0.2, 0.9) |
| Left atrial contractile strain | 0 (ref) | -0.8 (-1.8, 0.1) | -1.8 (-3.8, 0.2) |  | -0.7 (-1.2, -0.1) |
| **Spirits** | **0 drinks** | **≥ 1 drink/day** |  |  | **Per 1-drink increase** |
| Total LAVi | 0 (ref) | -1.6 (-3.1, -0.1) |  |  | -2.1 (-4.9, 0.7) |
| Left atrial reservoir strain | 0 (ref) | 0.5 (-0.9, 1.9) |  |  | -1.3 (-3.9, 1.2) |
| Left atrial conduit strain | 0 (ref) | 0.0 (-0.9, 1.0) |  |  | -0.4 (-2.2, 1.3) |
| Left atrial contractile strain | 0 (ref) | 0.5 (-0.5, 1.5) |  |  | -0.9 (-2.8, 0.9) |

LAVi, left atrial volume index.

All estimates are from model 2. Model 2 adjusts for age, sex, education, all alcohol types, marital status, smoking, physical activity, height, body mass index (BMI), systolic and diastolic blood pressure, diabetes, depression, diet adherence, and interaction of all covariates with time.

Supplemental Table 2. Mixed models estimates of each type of alcoholic beverage at baseline with change in LA measures from baseline to year 5.

| **Wine** | **0 drinks** | **1 drink/day** | **2-3 drinks/day** | **≥ 4 drinks/day** | **Per 1-drink increase** |
| --- | --- | --- | --- | --- | --- |
| Total LAVi | 0 (ref) | 0.0 (-0.4, 0.4) | 0.4 (-0.2, 1.0) | 0.1 (-0.4, 0.7) | 0.0 (-0.1, 0.1) |
| Left atrial reservoir strain | 0 (ref) | 0.0 (-0.4, 0.4) | -0.1 (-0.7, 0.4) | 0.4 (-0.1, 0.9) | 0.0 (0.0, 0.1) |
| Left atrial conduit strain | 0 (ref) | 0.0 (-0.3, 0.3) | -0.1 (-0.4, 0.3) | 0.2 (-0.2, 0.5) | 0.0 (0.0, 0.1) |
| Left atrial contractile strain | 0 (ref) | -0.1 (-0.3, 0.2) | -0.1 (-0.5, 0.2) | 0.2 (-0.2, 0.5) | 0.0 (0.0, 0.1) |
| **Beer** | **0 drinks** | **1 drink/day** | **≥ 2 drinks/day** |  | **Per 1-drink increase** |
| Total LAVi | 0 (ref) | -0.2 (-0.6, 0.1) | -0.3 (-1.0, 0.4) |  | 0.0 (-0.2, 0.3) |
| Left atrial reservoir strain | 0 (ref) | -0.1 (-0.4, 0.2) | -0.5 (-1.2, 0.2) |  | -0.1 (-0.3, 0.1) |
| Left atrial conduit strain | 0 (ref) | -0.2 (-0.4, 0.0) | -0.4 (-0.9, 0.1) |  | -0.1 (-0.2, 0.1) |
| Left atrial contractile strain | 0 (ref) | 0.1 (-0.1, 0.3) | -0.1 (-0.6, 0.4) |  | 0.0 (-0.2, 0.1) |
| **Spirits** | **0 drinks** | **≥ 1 drink/day** |  |  | **Per 1-drink increase** |
| Total LAVi | 0 (ref) | -0.1 (-0.4, 0.3) |  |  | 0.0 (-0.7, 0.7) |
| Left atrial reservoir strain | 0 (ref) | 0.2 (-0.2, 0.5) |  |  | 1.0 (0.4, 1.7) |
| Left atrial conduit strain | 0 (ref) | 0.2 (-0.1, 0.4) |  |  | 0.5 (0.0, 0.9) |
| Left atrial contractile strain | 0 (ref) | 0.0 (-0.2, 0.3) |  |  | 0.6 (0.1, 1.1) |

LAVi, left atrial volume index.

All estimates are from model 3. Model 3 adjusts for age, sex, education, intervention group, all alcohol types, marital status, smoking, physical activity, height, body mass index (BMI), systolic and diastolic blood pressure, diabetes, depression, diet adherence, interaction of all covariates with time, and differences from baseline to year 5 for the following: smoking, physical activity, BMI, systolic and diastolic blood pressure, diabetes, depression, diet adherence.

Supplemental Table 3. Multiple linear regression estimates of overall alcohol consumption with LA measures at baseline, by sex.

|  | **0 drinks** | **1 drink/day** | **2 drinks/day** | **> 2 drinks/day** | **Per 1-drink increase** |
| --- | --- | --- | --- | --- | --- |
| **Male = 301** | | | | | |
| Total LAVi | 0 (Ref.) | 1.0 (-1.8, 3.8) | 0.4 (-2.5, 3.4) | 3.7 (0.6, 6.8) | 0.8 (0.3, 1.3) |
| Left atrial reservoir strain | 0 (Ref.) | -0.9 (-3.4, 1.7) | -0.2 (-2.9, 2.4) | -1.8 (-4.6, 1.1) | -0.3 (-0.8, 0.1) |
| Left atrial conduit strain | 0 (Ref.) | 0.8 (-0.8, 2.5) | 1.4 (-0.3, 3.1) | 0.6 (-1.2, 2.4) | 0.0 (-0.3, 0.3) |
| Left atrial contractile strain | 0 (Ref.) | -1.6 (-3.5, 0.3) | -1.7 (-3.7, 0.3) | -2.3 (-4.4, -0.2) | -0.3 (-0.7, 0.0) |
| **Female = 202** | | | | | |
| Total LAVi | 0 (Ref.) | -0.5 (-2.6, 1.7) | -1.1 (-5.0, 2.9) | 1.9 (-6.7, 10.5) | 0.5 (-0.9, 1.9) |
| Left atrial reservoir strain | 0 (Ref.) | 0.2 (-1.7, 2.1) | -3.8 (-7.3, -0.2) | 5.3 (-2.4, 12.9) | -0.7 (-2.0, 0.6) |
| Left atrial conduit strain | 0 (Ref.) | 0.6 (-0.8, 2.0) | -1.0 (-3.6, 1.6) | 4.9 (-0.7, 10.5) | 0.0 (-0.9, 1.0) |
| Left atrial contractile strain | 0 (Ref.) | -0.3 (-1.5, 1.0) | -2.9 (-5.3, -0.4) | 0.7 (-4.5, 6.0) | -0.7 (-1.6, 0.1) |
| *Sex interactions* | | | | | |
| LAVi | | P = 0.76 | | | |
| Left atrial reservoir strain | | P = 0.59 | | | |
| Left atrial conduit strain | | P = 0.52 | | | |
| Left atrial contractile strain | | P = 0.20 | | | |

LAVi, left atrial volume index.

All estimates are from model 2. Model 2 adjusts for age, sex, education, all alcohol types, marital status, smoking, physical activity, height, body mass index (BMI), systolic and diastolic blood pressure, diabetes, depression, diet adherence, and interaction of all covariates with time.

Supplemental Table 4. Multiple linear regression estimates of overall alcohol consumption at baseline with change in LA measures from baseline to year 5, by sex.

|  | **0 drinks** | **1 drink/day** | **2 drinks/day** | **> 2 drinks/day** | **Per 1-drink increase** |
| --- | --- | --- | --- | --- | --- |
| **Male = 301** | | | | | |
| Total LAVi | 0 (Ref.) | 0.3 (-0.4, 1.0) | 0.6 (-0.2, 1.3) | 0.3 (-0.5, 1.1) | 0.0 (-0.1, 0.1) |
| Left atrial reservoir strain | 0 (Ref.) | 0.2 (-0.6, 1.0) | 0.0 (-0.8, 0.8) | 0.6 (-0.2, 1.4) | 0.1 (0.0, 0.2) |
| Left atrial conduit strain | 0 (Ref.) | -0.1 (-0.6, 0.4) | -0.2 (-0.7, 0.3) | 0.2 (-0.3, 0.8) | 0.1 (0.0, 0.2) |
| Left atrial contractile strain | 0 (Ref.) | 0.3 (-0.2, 0.8) | 0.3 (-0.3, 0.8) | 0.4 (-0.2, 1.0) | 0.1 (0.0, 0.1) |
| **Female = 202** | | | | | |
| Total LAVi | 0 (Ref.) | -0.4 (-1.0, 0.1) | 0.4 (-0.7, 1.4) | -0.2 (-2.9, 2.4) | 0.1 (-0.3, 0.5) |
| Left atrial reservoir strain | 0 (Ref.) | -0.2 (-0.6, 0.3) | -0.1 (-1.0, 0.7) | -1.8 (-3.9, 0.4) | -0.2 (-0.5, 0.1) |
| Left atrial conduit strain | 0 (Ref.) | -0.2 (-0.6, 0.1) | -0.1 (-0.7, 0.5) | -1.6 (-3.2, 0.0) | -0.1 (-0.4, 0.1) |
| Left atrial contractile strain | 0 (Ref.) | 0.0 (-0.3, 0.4) | 0.0 (-0.7, 0.6) | -0.6 (-2.2, 1.1) | -0.1 (-0.3, 0.1) |
| *Sex interactions* | | | | | |
| LAVi | | P = 0.71 | | | |
| Left atrial reservoir strain | | P = 0.38 | | | |
| Left atrial conduit strain | | P = 0.19 | | | |
| Left atrial contractile strain | | P = 0.92 | | | |

LAVi, left atrial volume index.

All estimates are from model 3. Model 3 adjusts for age, sex, education, intervention group, all alcohol types, marital status, smoking, physical activity, height, body mass index (BMI), systolic and diastolic blood pressure, diabetes, depression, diet adherence, interaction of all covariates with time, and differences from baseline to year 5 for the following: smoking, physical activity, BMI, systolic and diastolic blood pressure, diabetes, depression, diet adherence.

Supplemental Table 5. Mixed models estimates of change in overall alcohol consumption from baseline to year 5 with change in LA measures from baseline to year 5 (2 change categories).

| **Baseline alcohol consumption** | **Year 5** | |
| --- | --- | --- |
|  | 0-1 drinks/day | 2 or more drinks/day |
| LAVi | | |
| 0-1 drinks/day |  |  |
|  | 257 | 31 |
|  | 0 (Ref.) | 0.7 (0.1, 1.3) |
| 2 or more drinks/day |  |  |
|  | 38 | 125 |
|  | -0.1 (-0.8, 0.5) | 0 (Ref.) |
| *Left atrial reservoir strain* | | |
| 0-1 drinks/day |  |  |
|  | 244 | 31 |
|  | 0 (Ref.) | -0.6 (-1.2, 0.0) |
| 2 or more drinks/day |  |  |
|  | 37 | 122 |
|  | -0.3 (-1.0, 0.3) | 0 (Ref.) |
| *Left atrial conduit strain* | | |
| 0-1 drinks/day |  |  |
|  | 257 | 31 |
|  | 0 (Ref.) | -0.2 (-0.6, 0.2) |
| 2 or more drinks/day |  |  |
|  | 38 | 125 |
|  | -0.2 (-0.6, 0.2) | 0 (Ref.) |
| *Left atrial contractile strain* | | |
| 0-1 drinks/day |  |  |
|  | 257 | 31 |
|  | 0 (Ref.) | -0.4 (-0.8, 0.0) |
| 2 or more drinks/day |  |  |
|  | 38 | 125 |
|  | -0.1 (-0.6, 0.3) | 0 (Ref.) |

LAVi, left atrial volume index.

All estimates are from model 3. Model 3 adjusts for age, sex, education, intervention group, marital status, smoking, physical activity, height, body mass index (BMI), systolic and diastolic blood pressure, diabetes, depression, diet adherence, interaction of all covariates with time, and differences from baseline to year 5 for the following: smoking, physical activity, BMI, systolic and diastolic blood pressure, diabetes, depression, diet adherence.
